# Cardiovascular damage in parents of patients with mucopolysaccharidoses

**DOI:** 10.1101/2024.04.27.24306397

**Authors:** T.L. Nguyen, E.V. Reznik, A.N. Semyachkina, V.Yu. Voinova, M.A. Shkolnikova

**Author notes:** (corresponding author), postal code: 113000. Nguyen Thanh Luan, 108 Military Central Hospital, Tran Hung Dao Street 1, Hai Ba Trung, Hanoi, Vietnam. Reznik Elena Vladimirovna, Department of Propedeutics of Internal Diseases of the medical faculty of the Pirogov Russian national research medical University of the Ministry of healthcare of the Russian Federation, Moscow; Cardiologist of the GBUZ “City Clinical Hospital №31” of Healthcare Department of Moscow, Lobachevsky street, 42; https://elibrary.ru/authors.asp SPIN code 3494-9080; ResearcherIDN-6856-2016. Semyachkina Alla Nikolaevna — Department of Clinical Genetics, Veltischev Research and Clinical Institute for Pediatrics of the Pirogov Russian National Research Medical University, 125412, Moscow, st. Taldomskaya, 2. Voinova Victoria Yurievna – Department of Neuro- and Pathopsychology of Development, Faculty of Clinical and Special Psychology; Leading Researcher, Department of Clinical Genetics, Pirogov Russian National Research Medical University, Moscow, Ostrovityanova Street 1,. ResearcherID: O-3107-2013, Scopus: 9435408600. Shkolnikova Maria Alexandrovna — Children’s Scientific and Practical Center for Heart Rhythm Disorders of the Ministry of Health of the Russian Federation, Pediatric Cardiologist of Pirogov Russian National Research Medical University, 125412, Moscow, st. Taldomskaya, 2.

## Abstract

Mucopolysaccharidoses (MPS) are a group of lysosomal storage diseases. Cardiovascular pathology occurs in all types of MPS, represented by valvular defects, myocardial hypertrophy, and coronary artery disease. Cardiovascular abnormalities in parents of carriers with MPS are poorly understood, which was the purpose of our work.

In 2022 year 21 parents (81% female) of children with MPS were examined.The median (25th and 75th percentiles) of age was 36 (33; 37) years. All MPS carriers-parents underwent a standard clinical and laboratory examination, ECG, echocardiography, 24-hour Holter ECG monitoring.

There were no confirmed myocardial and brain infarctions, diabetes mellitus in the examined carriers. Arterial hypertension was diagnosed in 14.3% of carriers. A decrease in LV ejection fraction <40% was found in 4.8%, up to 40-50% in 9.5% of carriers. LV wall thickness ≥1.5 cm was detected in 66.7% of carriers, asymmetric LVH in 85.7%. Thickening of the mitral valve leaflets (MV) was detected in 76.2% of carriers. Hydropericardium was detected in 23.8% of carriers. Atrial fibrillation was registered in 4.8%, atrial flutter - in 4.8%, paroxysmal supraventricular tachycardia - in 33.3%, sinus bradycardia - in 14.3%; conduction disorders - in 71.4% of carriers. A short PR interval was detected on the ECG 9.5% of carriers. A prolonged QT interval was registered in 14.3% of carriers, transient ST-segment depression in 47.6%, ST-segment elevation in 14.3% of carriers.

Our results suggest the possibility of clinical manifestations of cardiac involvement in MPS carriers.

## Introduction

Mucopolysaccharidoses (MPS) are a group of inherited metabolic diseases caused by deficiencies in enzymes involved in the sequential degradation of glycosaminoglycans leading to substrate accumulation in various tissues and organs [1]. Currently 13 types of MPS have been described, including 2 types discovered in the 2020s [2]. All MPSs are inherited in an autosomal recessive manner except MPS II, which is X-linked [2]. The incidence of MPSs varies within each disorder and in different populations and ethnic groups, with a prevalence of all their types of about 1 per 40,000—50,000 live births [3]. One of the most important clinical aspects of MPSs is cardiovascular pathology. Cardiac abnormalities occur in all types of MPS, the most common being valvular defects and cardiac hypertrophy, which result from the accumulation of glycosaminoglycans in the spongiosis of the heart valves, myointima of coronary arteries and myocardium [4]. Cardiovascular abnormalities in the parents of patients with MPS are little studied, which became the purpose of our work.

## Material and methods

In 2022 year 21 parents (17 women, 81.0%) of children with confirmed MPS were examined: 2 (9.5%) - type I, 6 (28.6%) - type II, 1 (4, 8%) - type III, 11 (52.4%) - type IV, 1 (4.8%) - type VI. The median (25th and 75th percentiles are indicated in parentheses) age was 36 (33; 37) years. All patients underwent a standard clinical and laboratory examination, 12-lead electrocardiography (ECG), echocardiography, 24-hour Holter ECG monitoring (Table 1).

**Table 1.**
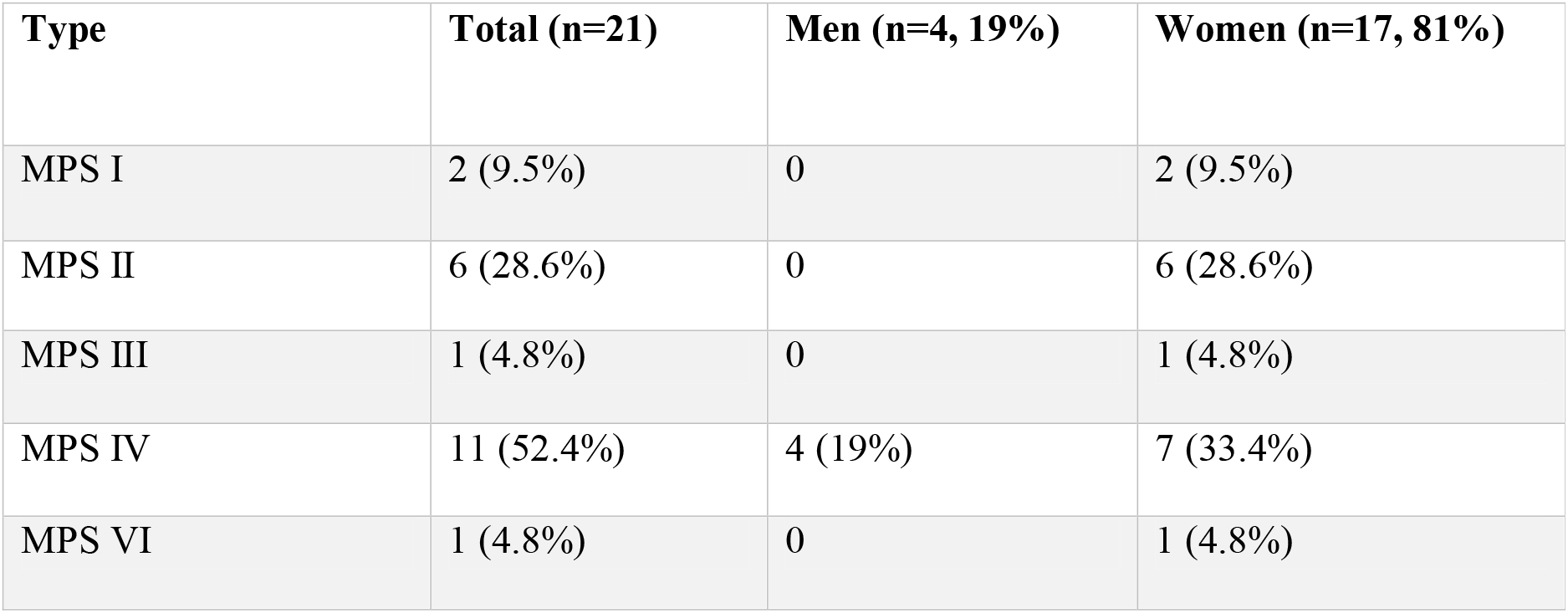
Characteristics of carriers of mucopolysaccharidoses

Due to small sample size the distribution of the studied quantitative characteristics differed from the normal one, non-parametric criteria were used in statistical calculations and quantitative indicators are presented as a median and an interquartile range. Differences between nominal variables were compared using a chi-square test. Fisher’s exact test was used when more than 20% of cells have expected frequencies less than 5. P value <0.05 was considered statistically significant. The analysis was performed by a biostatistician using the statistical software SPSS (version 26.0; SPSS Institute, USA) and STATISTICA (version 12.0; StatSoft, USA).

## Results

There were no confirmed myocardial and brain infarctions, diabetes mellitus in the examined subjects. Arterial hypertension was diagnosed in 3 cases (14.3%: in 2 - 1 degree, in 1 - 2 degree). congenital heart disease up to 24 years (open ductus arteriosus and open foramen ovale) - in 1 (4.8%), chronic pyelonephritis - in 2 (9.5%) cases.

According to 12-lead ECG and 24-hour Holter ECG monitoring, paroxysmal atrial fibrillation was registered in 1 (4.8%) carrier, paroxysmal atrial flutter - in 1 (4.8%), unstable paroxysmal supraventricular tachycardia - in 7 (33.3%), sinus bradycardia - in 3 (14.3%), sinus tachycardia - in 6 (28.6%) carriers (Figure 1).

**Figure 1.**
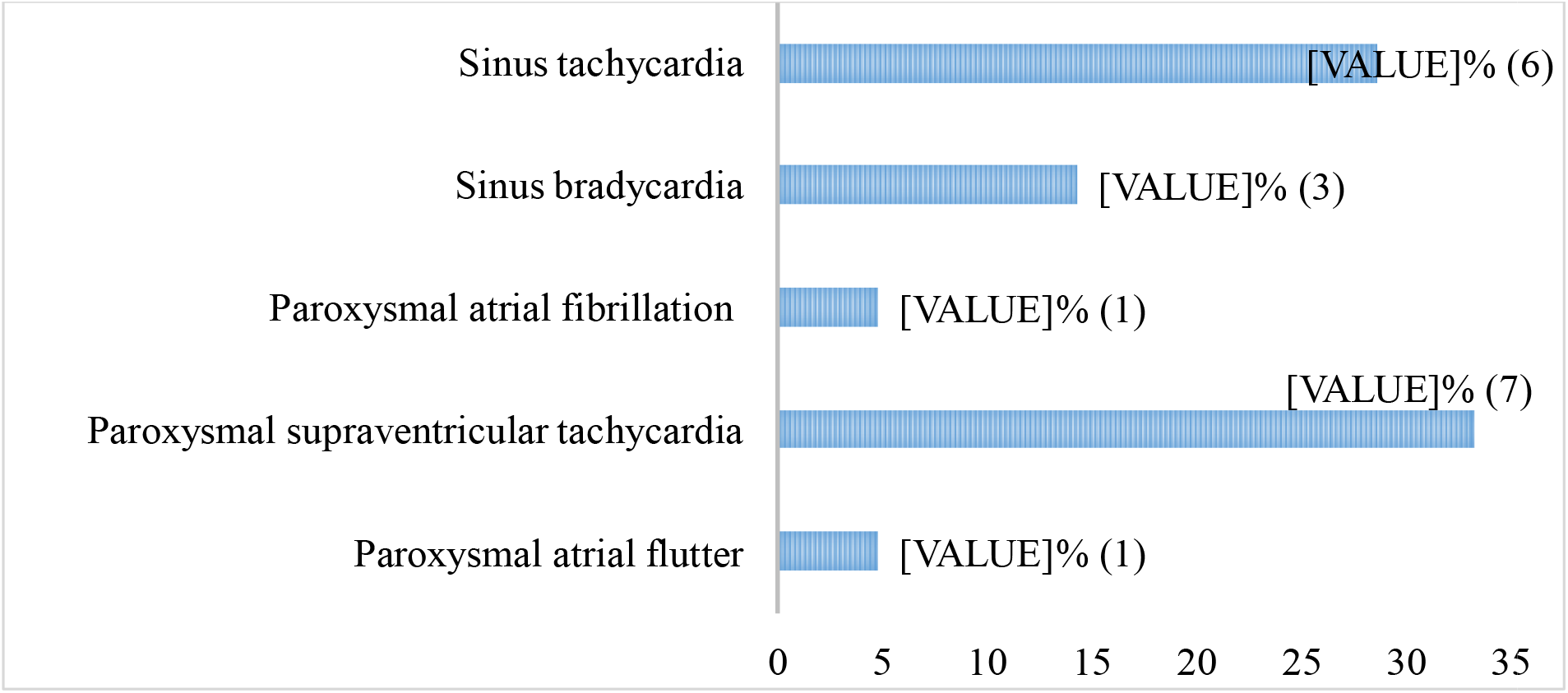
Rhythm disturbances in MPS carriers

Conduction disorders were detected in 15 (71.4%) carriers, of which first-degree atrioventricular block - in 2 (9.5%), bundle branch block (right and / or left) - in 5 (23.8%), nonspecific intraventricular conduction delay - in 9 (42.9%) carriers (Figure 2).

**Figure 2.**
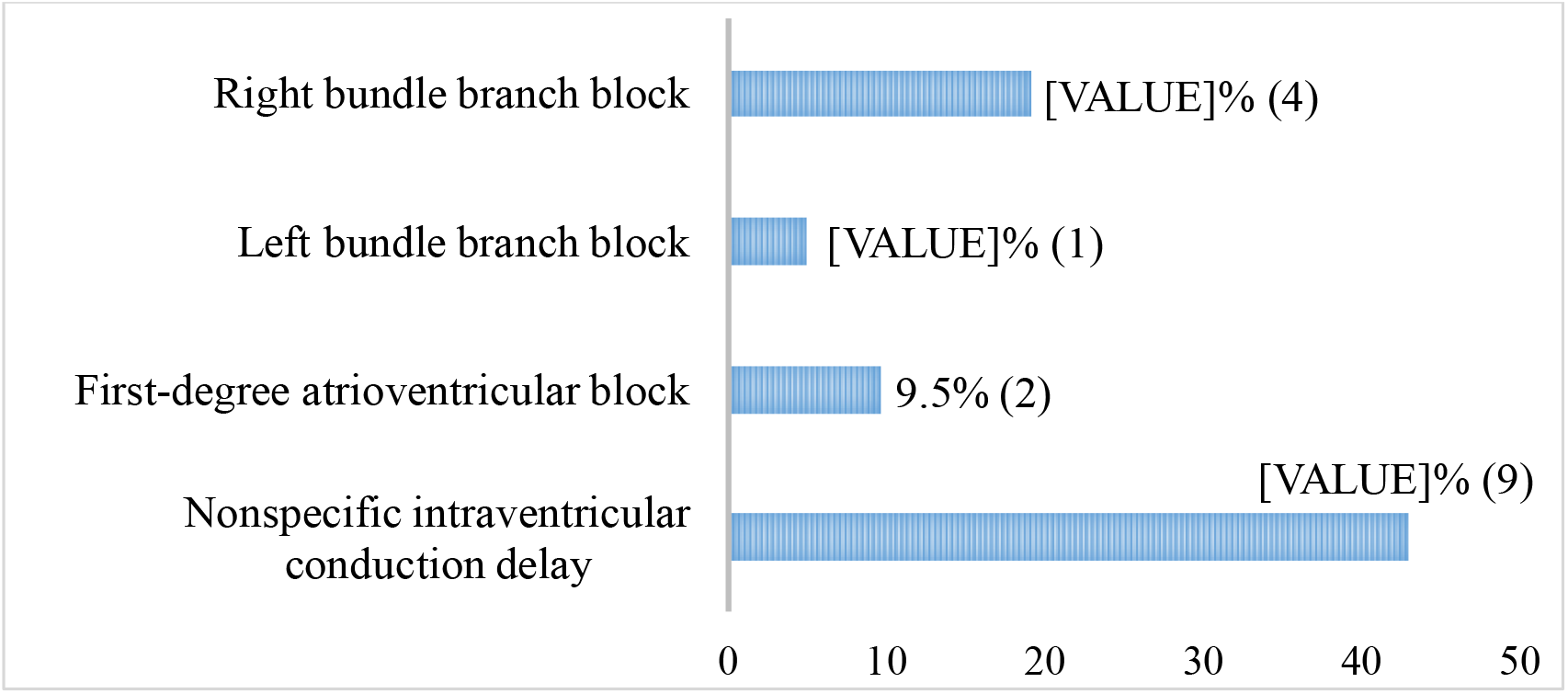
Conduction disorders in MPS carriers

Low voltage of QRS complex (zenith-to-nadir QRS amplitudes of the QRS complexes of less than 0.5 mV in all the frontal leads and/or less than 1.0 mV in all the precordial leads) was found in 4 (19%) cases, Sokolow-Lyon ECG criteria for LVH were found in 2 (9.5%) cases (Figure 3).

**Figure 3.**
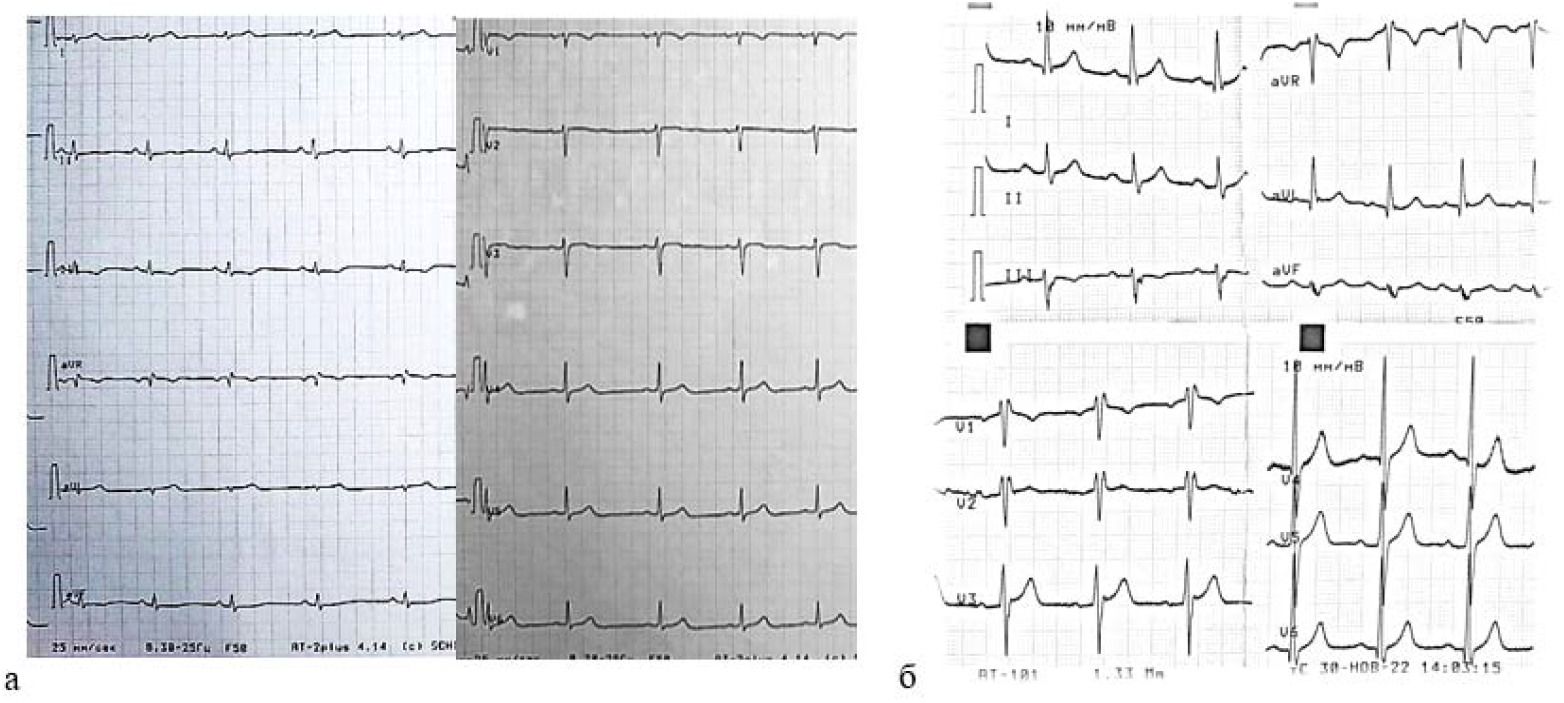
Voltage changes of the QRS complex on MPS carriers ECG. Low voltage of QRS complex (a) and left ventricular hypertrophy voltage criteria (b) (speed 25 mm/s)

The presence of pathologic Q wave on ECG was registered in 12 (57.2%) cases (all carriers denied a history of myocardial infarction): in 11 (52.4%) cases - in the inferior leads, in 1 (4.8%) case - in the lateral leads. Poor R wave progression was detected in 4 (19%) cases (Figure 4).

**Figure 4.**
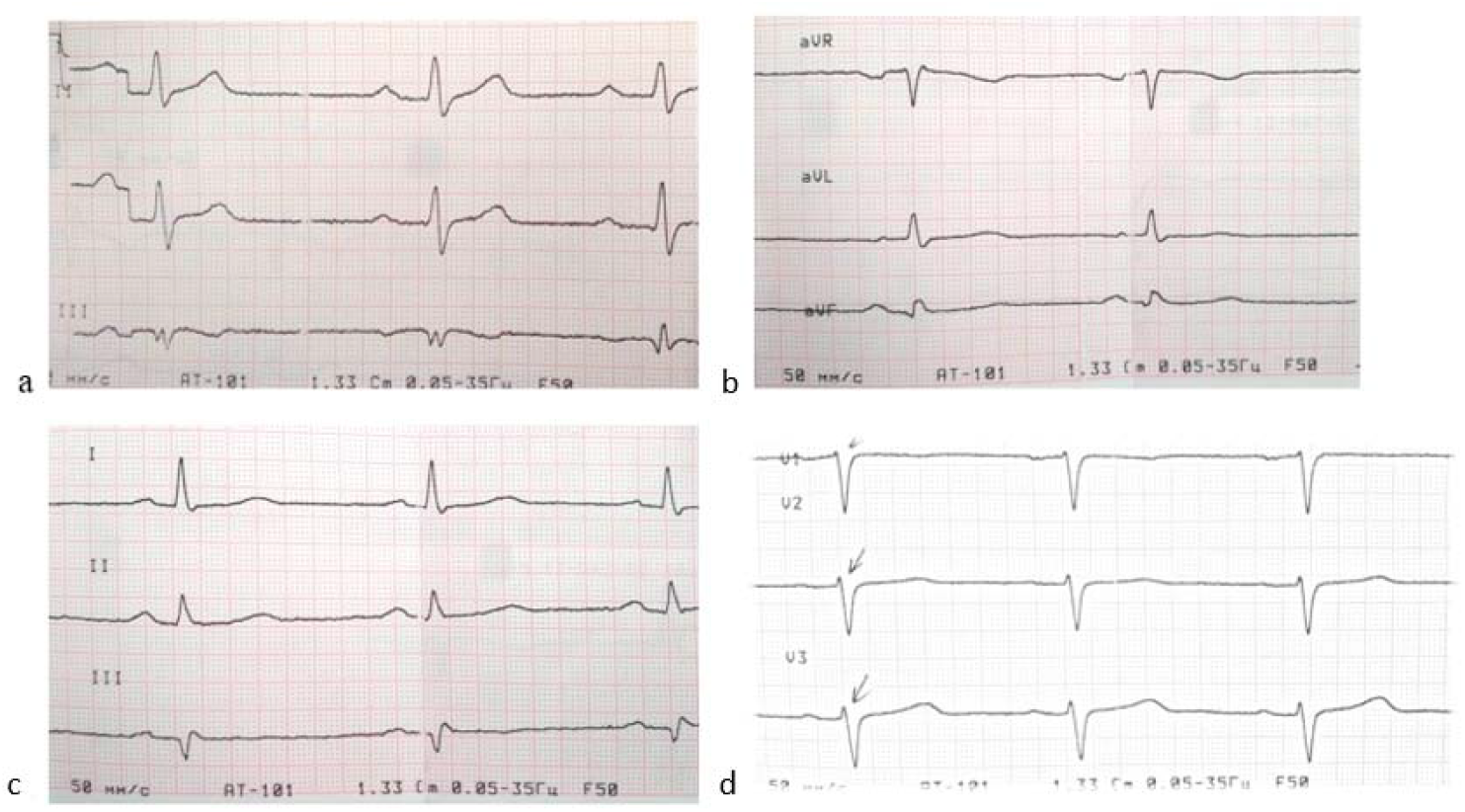
The presence of a Q wave on the ECG in the inferior leads (a, b, c) and poor R wave progression (d) in MPS carriers (speed 50 mm/s).

Nonspecific ventricular repolarization abnormalities were detected in 6 (28.6%) cases, early repolarization pattern - in 3 (14.3%) cases. A short PR interval (of less than 120 ms) was detected on the ECG in 2 (9.5%) cases. A prolonged QT interval was registered in 3 (14.3%) cases. Transient ST-segment depression was detected in 10 (47.6%) cases, ST-segment elevation - in 3 (14.3%) cases, all of which were registered during exercise or tachycardia.

According to echocardiographic examinations, the ejection fraction (EF) of the left ventricle was 60 (54; 62)%. LVEF<40% was detected in 1 (4.8%) case, up to 40-50% - in 2 (9.5%) cases. 85.7% of cases had preserved LVEF.

LV hypertrophy (LVH) was detected in 20 (95.2%) carriers, LV wall thickness greater than 1.5 cm was found in 14 (66.7%) carriers. Symmetric LVH was detected in 3 (14.3%) carriers, apical - in 11 (52.4%, isolated apical LVH - in 5 (23.8%)), papillary muscle hypertrophy - in 11 (52.4%, isolated - in 3 (14.3%) carriers), interventricular septal hypertrophy - in 4 (19%) carriers, lateral - in 1 (4.8%) in combination with apical LVH, isolated hypertrophy of the posterior LV wall - in 1 (4.8%) carrier. No LV outflow tract obstruction was identified (Figure 5, 6).

**Figure 5.**
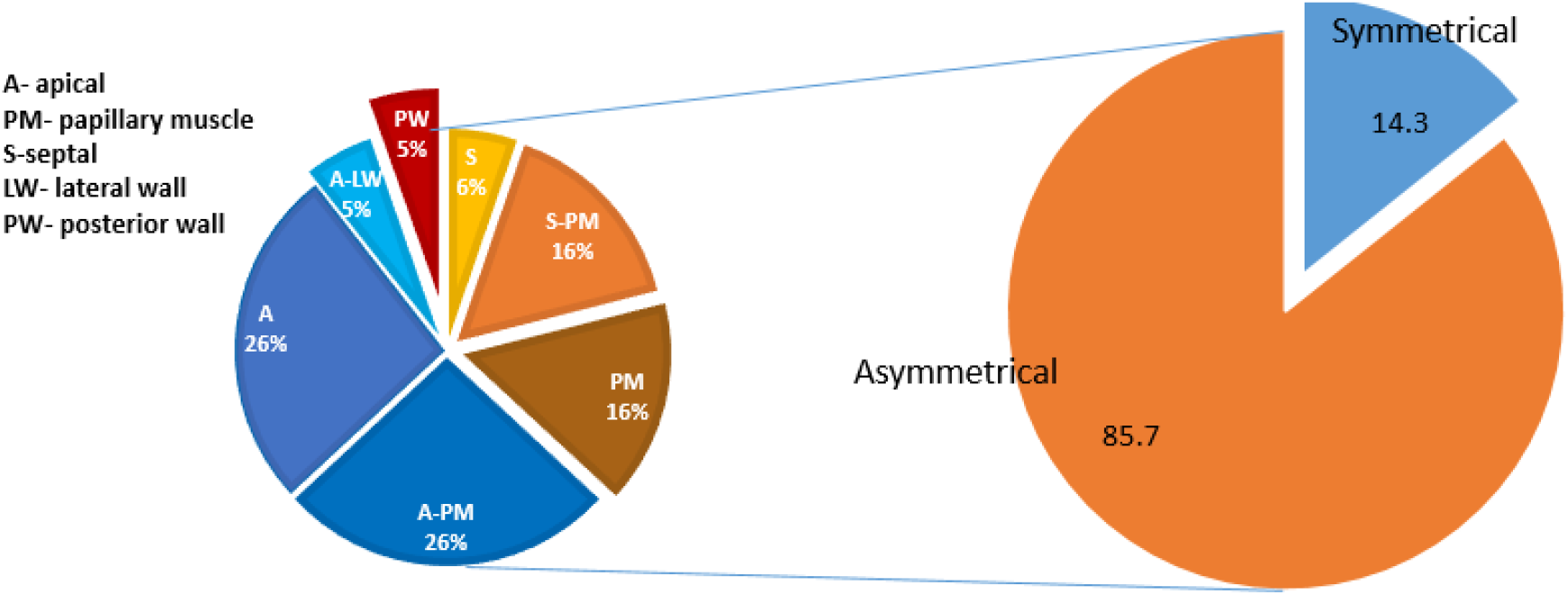
Characteristics of left ventricular myocardial hypertrophy in MPS carriers

**Figure 6.**
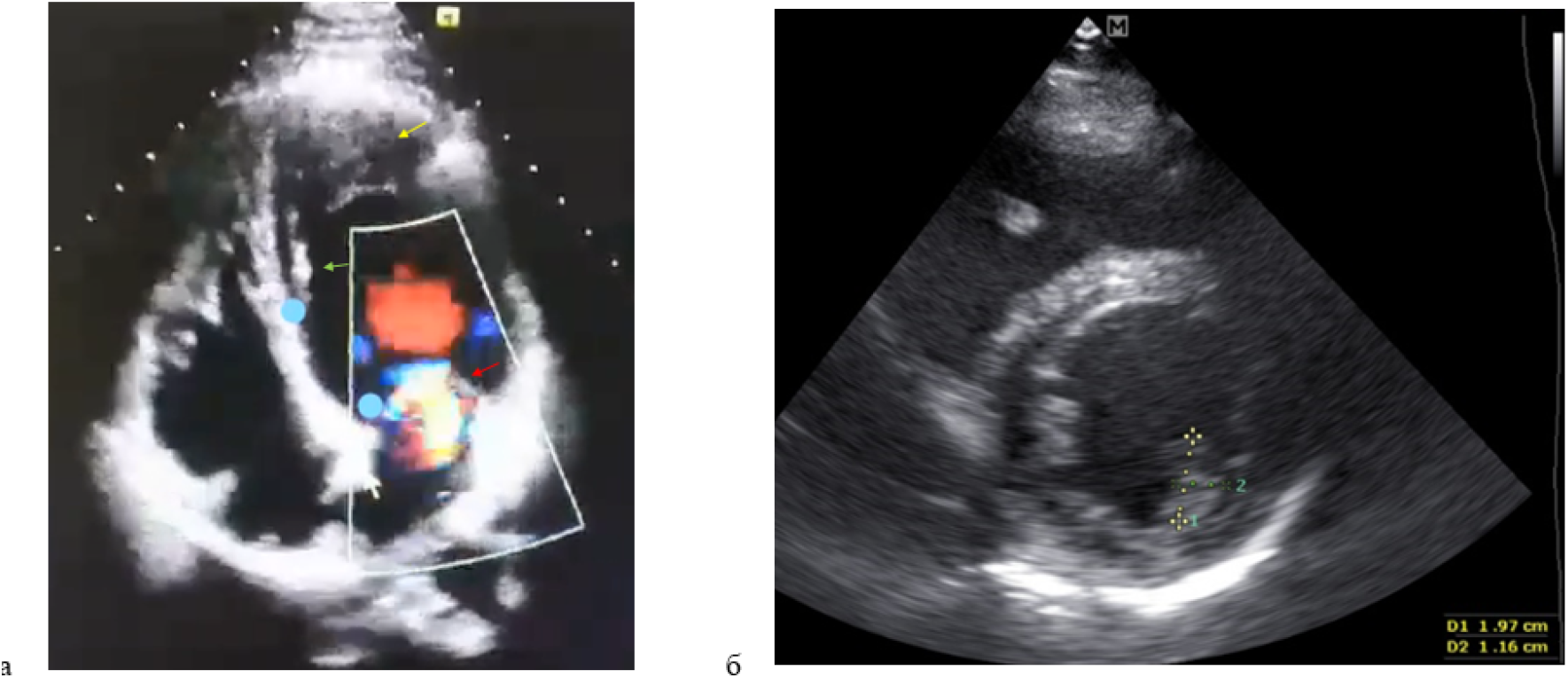
Echocardiography of the heart in a heterozygous female carrier of MPS type I (a) with severe left ventricular apex hypertrophy (yellow arrow), thickening of the mitral valve leaflets, moderate mitral regurgitation (red arrow), thickening of the atrial septum (white arrow), dilatation of the right ventricular cavity, hyperechogenicity endocardium (green arrow); and in a heterozygous male carrier of MPS IV (b) with left ventricular papillary muscle hypertrophy.

Prolapse of the anterior leaflet of the mitral valve was detected in 7 (33.3%) carriers. Thickening of the mitral valve leaflets was detected in 16 (76.2%) cases, mild (4-5 mm) - in 3 (14.3%) cases, moderate (5-7 mm with thickening of the mitral valve chords) - in 9 (42.9%), severe (>7 mm with thickening of the chords and fibrosis of the apex of the papillary muscle) - in 4 (19%) carriers (Figure 7). Hydropericardium was detected in 5 (23.8%) cases.

**Figure 7.**
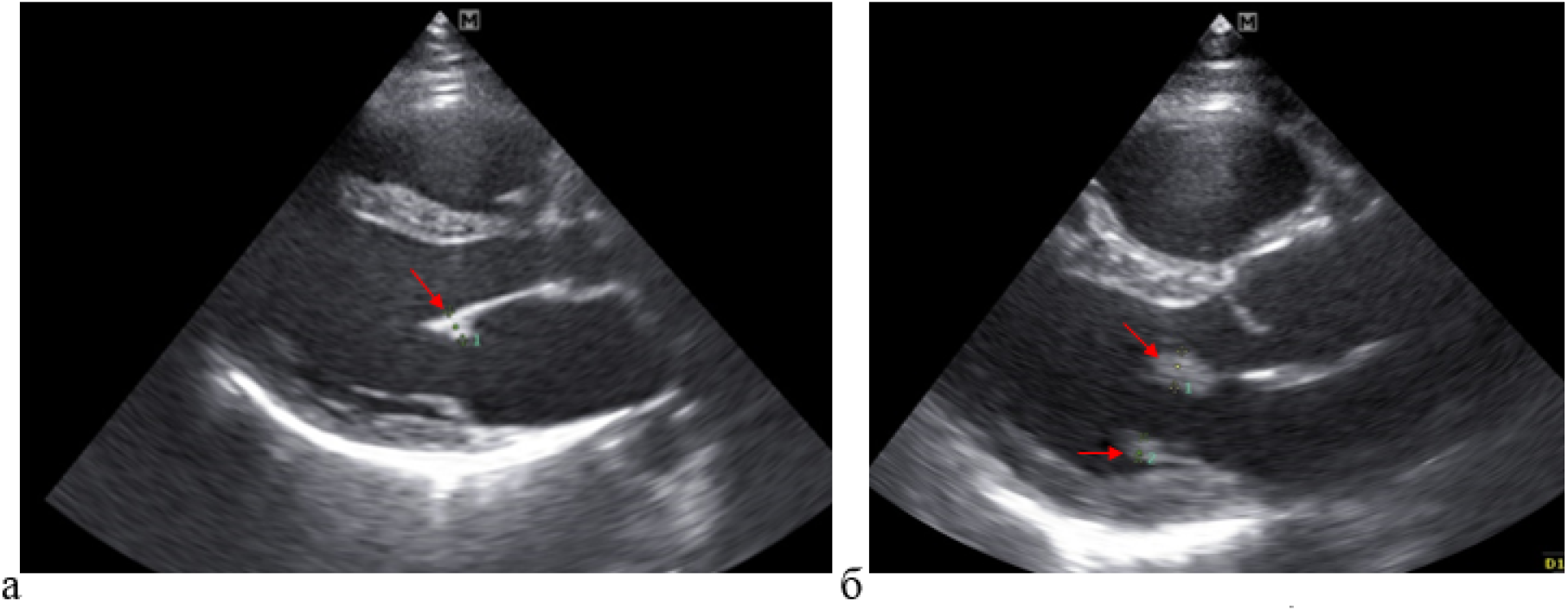
Thickening of the mitral valve leaflets on echocardiography: a) Thickening (6 mm) of the mitral valve anterior leaflet and shortening of mitral valve posterior leaflet in young MPS 4a type mutation carrier; b) Thickening of the mitral valve anterior (7 mm) and posterior (4,5 mm) leaflets in young MPS 4a type mutation carrier.

Mitral regurgitation was detected in 20 (95.2%) carriers, with I degree - in 12 (57.2%), II - in 6 (28.6%), III - in 2 (9.5%) carriers. Tricuspid regurgitation was detected in 20 (95.2%) carriers, with grade I in 10 (47.6%), grade II in 8 (38.1%), grade III in 2 (9.5%) carriers. Aortic regurgitation - in 4 (19%) carriers, with I degree - in 2 (9.5%), II - in 2 (9.5%) carriers. Pulmonary regurgitation was detected in 7 (33.3%) carriers, with I degree - in 3 (14.3%), II - in 4 (19%) carriers (Figure 8).

**Figure 8.**
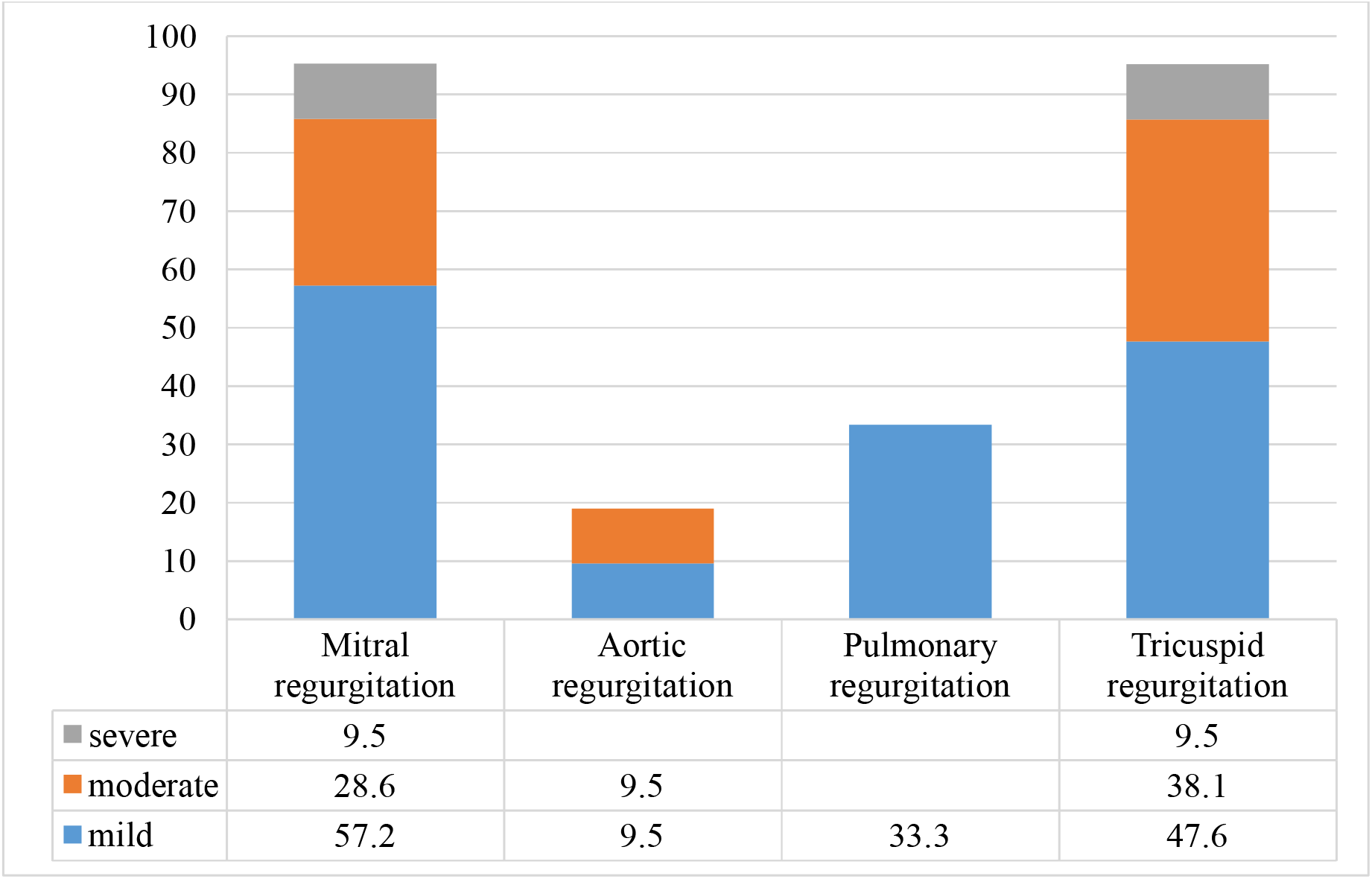
Valvular regurgitation in MPS carriers

LV end-diastolic volume, LV end-systolic volume, LV myocardial mass, wall thickness of the left and right ventricle were significantly higher, however LVEF was lower in male than in female MPS carriers (Table 2).

**Table 2.** Comparative characteristics of echocardiographic parameters for male and female MPS carriers *Abbreviation: CHF - chronic heart failure, LV - left ventricle*

| Parameter | Male (n=4) | Female (n=17) | P |
| --- | --- | --- | --- |
| LV wall thickness, mm | 18.6 (17.2; 21.2) | 14 (12.4; 15) | 0.024 |
| The thickness of the wall of the pancreas, mm | 4.7 (4.3; 5.5) | 3.6 (3.45; 3.9) | 0.007 |
| Mass of LV myocardium, g | 212.5 (172; 292.5) | 150 (138; 174) | 0.04 |
| LV ejection fraction, % | 43.25 (39.5; 54.8) | 61 (55.7; 62) | <0.001 |
| End-diastolic LV volume, ml | 109.4 (100.1; 133.5) | 89.7 (78.9; 98) | 0.024 |
| LV end-systolic volume, ml | 60.3 (54.35; 66) | 35.7 (33.3; 43.1) | 0.004 |

According to laboratory tests, the level of NT-proBNP>125 pg/ml was detected in 5 (23.8%) cases, the level of troponin I was within normal limits (≤0.01 ng/ml) in all cases.

## Discussion

In mutant MPS gene carriers changes in enzyme activity and properties have been observed and reported. They have lower levels of enzymatic activity, but overlap with those of normal people [5]. Studies of fibroblast, plasma and leukocyte samples show differential biochemical behavior of alpha-L-iduronidase derived from carriers of the mutant MPS gene compared to normal individuals [5–7]. There is evidence of possible progressive clinical manifestations in carriers of MPS II [8]. However, the results of studies of heart damage in mutant MPS gene carriers have not yet been published.

The results of our study indicate the presence of cardiovascular abnormalities in mutant MPS gene carriers. A relatively high proportion of subjects were found to have valvular lesions, including structural valvular changes (cusp thickening, mitral valve prolapse), valvular dysfunction (regurgitation); as well as thickening of the heart wall, not due to pre- and afterload factors. Decreased LVEF (<50%) was detected in 14.3% of cases, increased NT-proBNP level (>125 pg/ml) was detected in 23.8% of cases, while the average age of the study participants was 36 years, which indicates the need for screening and early assessment of the function of the cardiovascular system in such individuals.

In addition, such ECG changes as signs of LV myocardial hypertrophy, low voltage of QRS complexes, arrhythmias and conduction disturbances were observed with a relatively high frequency, proving there were abnormalities of cardiac electrophysiology in such individuals, requiring further study.

## Conclusion

For the first time, cardiovascular system was assessed in parents of patients with MPS. Our results indicate the possibility of clinical manifestations of heart disease in MPS carriers. Further comparative studies are required in larger populations to assess the rate of progression of the identified abnormalities and the effectiveness of drug therapy in MPS carriers.

## Data Availability

Study data can be provided via author's email upon request

## Author Contribution

All the authors contributed significantly to the study and the article, read and approved the final version of the article before publication

## Notes

### Competing Interest Statement

The authors have declared no competing interest.

### Funding Statement

No external funding was received

### Author Declarations

Ethics сommittee of the Pirogov Russian National Research Medical University gave ethical approval for this work

